# Respiratory Syncytial Virus (RSV) Hospital Admission Rates and Patients’ Characteristics Before the Age of Two in England, 2015-2019

**DOI:** 10.1101/2023.04.04.23288132

**Authors:** Maria João Fonseca, Saskia Hagenaars, Mathieu Bangert, Clare Flach, Richard Hudson

**Affiliations:** IQVIA, Lisbon, Portugal; IQVIA, London, United Kingdom; Sanofi, Lyon, France; Sanofi, Reading, United Kingdom

**Keywords:** RSV, respiratory syncytial virus, hospital admission, burden of disease, England

## Abstract

**Background:** A granular understanding of respiratory syncytial virus (RSV) burden in England is needed to prepare for new RSV prevention strategies. We estimated the rates of RSV hospital admissions in infants before age two in England and describe baseline characteristics.

**Methods:** A birth cohort of all infants born between 01/03/2015 and 28/02/2017 (n=449,591) was established using Clinical Practice Research Datalink-Hospital Episode Statistics. Case cohorts included infants with an admission for 1) RSV-coded, 2) bronchiolitis-coded, 3) any respiratory tract infection (RTI)-coded <24 months and 4) RSV-predicted by an algorithm <12 months. Baseline characteristics were described in case and comparative cohorts (infants without corresponding admission). Cumulative incidence and admission rates were calculated. Multiple linear regression was used to estimate the proportion of RTI healthcare visits attributable to RSV.

**Results:** The RSV-coded/RSV-predicted case cohorts were composed of 4,813/12,694 infants (cumulative incidence: 1.1%/2.8%). Case cohort infants were more likely to have low birth weight, comorbidities and to be born during RSV season than comparative cohort infants, yet >77% were term healthy infants and >54% born before the RSV season. During the first year of life, 11.6 RSV-coded and 34.4 RSV-predicted hospitalizations occurred per 1,000 person-years. Overall, >25% of unspecified lower RTI admissions were estimated to be due to RSV.

**Conclusions:** In England, one in 91 infants had an RSV-coded admission, likely underestimated by ∼3-fold. Most infants were term healthy infants born before the RSV season. To decrease the total burden of RSV at the population level, immunization programs need to protect all infants.

## Introduction

Respiratory syncytial virus (RSV) causes substantial burden on healthcare systems worldwide.^1^ Although most infants manifest mild symptoms manageable at home, the clinical manifestations of an RSV infection may range from a mild upper respiratory tract infection (URTI) to a severe lower respiratory tract infection (LRTI) and hospitalization.^2^ There is emerging evidence that it can also lead to long term respiratory morbidity.^3–5^

RSV is the main cause of acute LRTI in children globally.^1^ A recent systematic review estimated that RSV is responsible for 33 million episodes of LRTI, resulting in 3.6 million hospital admissions, around 26,300 in-hospital deaths, and >100,000 overall deaths per year in children aged <5 years worldwide.^1^ In England, between 2007 and 2017, 7,062 RSV-coded hospitalizations occurred annually in children aged <5 years, and, of those, 6,337 occurred in infants <1 year, corresponding to a rate of 8.6 per 1,000 infants.^6^

Likely due to lack of testing for RSV^7^, the estimates of RSV morbidity and mortality based on RSV diagnosis codes alone are most likely underestimated^6^ and other methods, such as statistical modeling^8,9^ or the use of proxies for RSV activity like bronchiolitis^10–12^, have been used. Recognizing this underestimation, Reeves et al. developed an algorithm which estimated that 33,561 RSV-associated hospitalizations occurred annually in children aged <5 years in England between 2007 and 2012, at a rate of 35.1 per 1,000 infants aged <1 year.^8^ This is ∼4 times higher than RSV diagnosis codes alone suggest.^6^ The same authors concluded that ∼78% of hospitalizations for bronchiolitis in children aged <5 years may be RSV related, and increases to 82% in infants <6 months.^8^

The likelihood of developing severe RSV infection is higher in infants with known risk factors such as chronic lung disease (CLD) of prematurity, congenital heart disease (CHD), neuromuscular disorders, immunodeficiencies, and pre-term birth.^13,14^ Infants born just before and during the RSV season, which occurs from October to February in England, also have a higher risk of serious RSV infection.^15^

This study aimed to estimate the rates of RSV hospital admissions in infants up to 24 months old in England, using different approaches, including proxies and statistical modeling, and describe the sociodemographic and clinical characteristics of infants with a hospitalization born before and during the season.

## Materials and Methods

### Data sources

This was a retrospective cohort study, using Clinical Practice Research Datalink (CPRD)- HES (Hospital Episode Statistics) linked data. CPRD is a longitudinal primary care electronic medical record data source, that covers 20% of the English population.^16^ HES contains details of all admissions to National Health Service (NHS) hospitals in England. HES data can be linked at patient-level for a subset of CPRD-contributing practices.

### Cohorts’ definitions

#### Diagnosis coded case cohorts

A birth cohort of all infants born between 01/03/2015 and 28/02/2017 within the CPRD-HES linked dataset was established (n=449,591). Three diagnosis coded case cohorts were defined, each included infants who had at least one hospital admission of interest up to 24 months of age, coded in HES with: 1) J12.1, J20.5, J21.0, or B97.4 (only included if the infant had also an RTI code in the same admission) - RSV-coded case cohort; 2) J21 - bronchiolitis-coded case cohort; and 3) J00-06, J12-J18, J20-22 - RTI-coded case cohort.

#### RSV-predicted case cohort

A complementary RSV-predicted case cohort, based on the Reeves at al.^9^ published algorithm, was defined, and included infants identified by the logistic regression prediction model defined below.

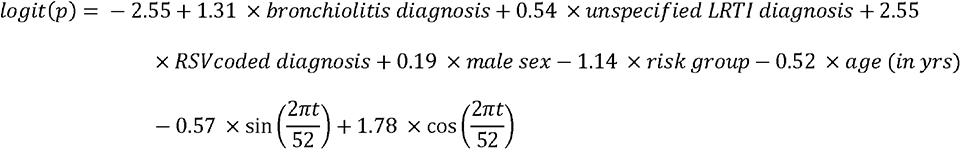

The algorithm was applied to all RTI admissions in order to predict if the admission was positive for RSV (probability ≥ 0.5) or not (probability < 0.5). Infants who had at least one RTI admission predicted to be due to RSV up to 12 months of age were included in the RSV- predicted case cohort.

#### Comparative cohorts

Each case cohort was compared with a comparative cohort composed of all infants who did not have the hospital admission of interest, according to the case cohort definition. For example, the RSV-coded case cohort was compared with the remaining sample not fitting in the RSV-coded case cohort (including infants belonging to the other case cohorts and infants not fitting in any case cohort).

Case and comparative cohorts were not mutually exclusive (Figure 1).

**Figure 1.**
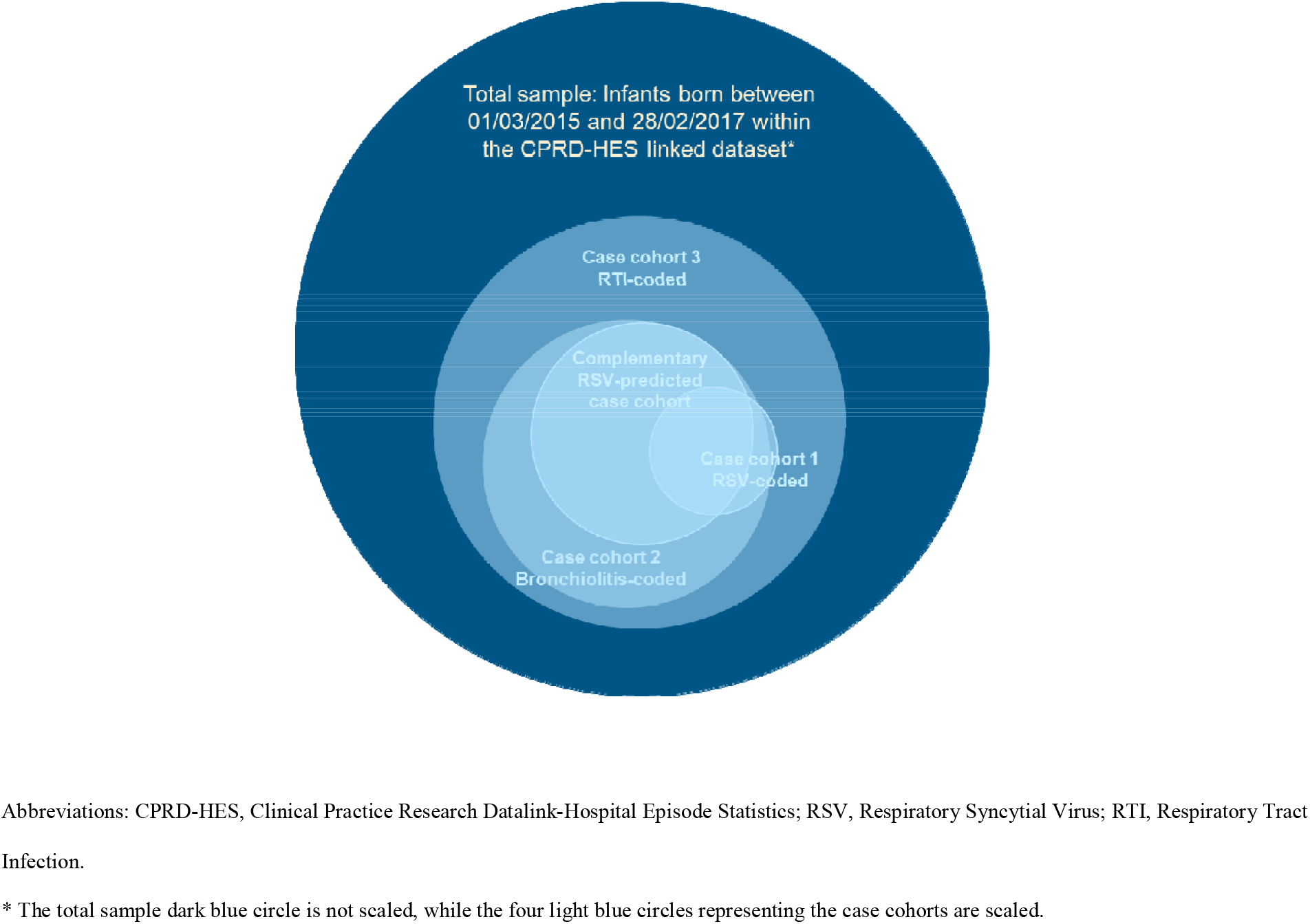
Diagrammatic Representation of the Case Cohorts Used in This Study.

### Variables’ definitions

Births between 1^st^ October to 28^th^/29^th^ February were considered during season, while those between 1^st^ March to 30^th^ September were considered before season. High-risk status was defined as a diagnosis of at least one of: CLD, CHD, prematurity, neurological disorders, or immunodeficiency. Palivizumab eligibility was defined according to UK guidelines^17^ as infants with at least one record of CLD, CHD or Severe Combined Immunodeficiency Syndrome.

### Statistical analysis

Statistical analysis was performed using SAS Enterprise Guide 8.2 and R version 3.5.2 software.

The cumulative incidence was calculated as the number of infants with at least one admission of interest divided by the total number of infants in the birth cohort. Sociodemographic and clinical characteristics were described for all cohorts, using numbers and proportions.

The total number of RSV-coded, bronchiolitis-coded, any RTI-coded and RSV-predicted admissions up to 24 and 12 months of life, that occurred among the birth cohort participants, was described, as well as the respective rates calculated as the total number of admissions divided by the total person-years at risk and multiplied by 1,000.

Multiple linear regression modeling was used to understand the total burden of RSV-associated hospital admissions and consultations during the study period (01/03/2015 - 28/02/2019) for infants aged <24 months old. Seasonality in pathogens that cause RTI were used to estimate the proportion of consultations and hospital admissions with unspecified etiology that were attributable to different pathogens using six multiple linear regression models for three outcomes of interest (number of weekly primary care consultations for RTI, unspecified LRTI hospitalizations, and unspecified URTI hospitalizations), separately for before and during season births. The predictors in each model were the number of hospitalizations per calendar week in the total cohort with a code attributed to each of the following nine pathogens: RSV, influenza, parainfluenza, rhinovirus, human metapneumovirus, *Bordetella* species, *Streptococcus pneumoniae*, *Haemophilus influenzae*, and *Mycoplasma pneumoniae*. The count of each outcome and each predictor were aggregated across the years of the study period to the corresponding week of the year. For each of the six models we determined the proportion of the outcome attributable to each pathogen^18^ by conducting a linear regression model of the outcome on all the predictors applying a backward covariate selection, selecting the model with the lowest Akaike Information Criterion. The best fitting model was selected.

## Results

### Case Cohorts and Cumulative Incidence

Of the 449,591 infants included in the birth cohort, 4,813 had at least one RSV-coded admission before 24 months of age, corresponding to a cumulative incidence of 1.1%. These infants formed the RSV-coded case cohort. Likewise, 22,913 infants had at least one bronchiolitis-coded admission (cumulative incidence: 5.1%), forming the bronchiolitis-coded case cohort, and 56,871 had at least one RTI-coded admission (cumulative incidence: 12.7%), forming the RTI-coded case cohort. The RSV-predicted case cohort with at least one RSV- predicted admission up to the age of 12 months included 12,694 infants (cumulative incidence: 2.8%). Of all infants included in the birth cohort, 392,720 (87%) had no RSV-, bronchiolitis- or any other RTI-coded admission and no RSV-predicted admission.

### Sociodemographic and Clinical Characteristics

Infants with an RSV admission were more likely to be male (55.6% vs. 51.1%), born in the south-west/south-central regions of England (32.3% vs. 22.5%) and White (77.1% vs. 66.0%), compared to infants with no RSV admission (Table 1). Among infants with an RSV admission, the proportion belonging to the most deprived quintile of Index of Multiple Deprivation (IMD) was higher than the proportion belonging to the least deprived quintile (27.0% vs. 17.2%). Although the distribution across the IMD quintiles was similar in the case and comparative cohorts, a slightly higher proportion of infants was found in the most deprived quintile of IMD among the RSV-coded case cohort (27.0% vs. 25.1%). Infants with an RSV admission were more likely to be born during the RSV season (45.7% vs 40.2%) and with low birth weight (5.1% vs. 1.7%), even though birth weight had a high degree of missingness. Comorbidities were more frequent among infants with an RSV admission compared to those with no admission, consequently they were more likely to have high-risk status (24.1% vs 6.8%) and to be eligible to receive palivizumab (2.5% vs. 0.27%). Even so, 76.9% of infants in the RSV-coded case cohort were palivizumab non-eligible term and 54.3% were born before the RSV season (Table 1).

**Table 1.**
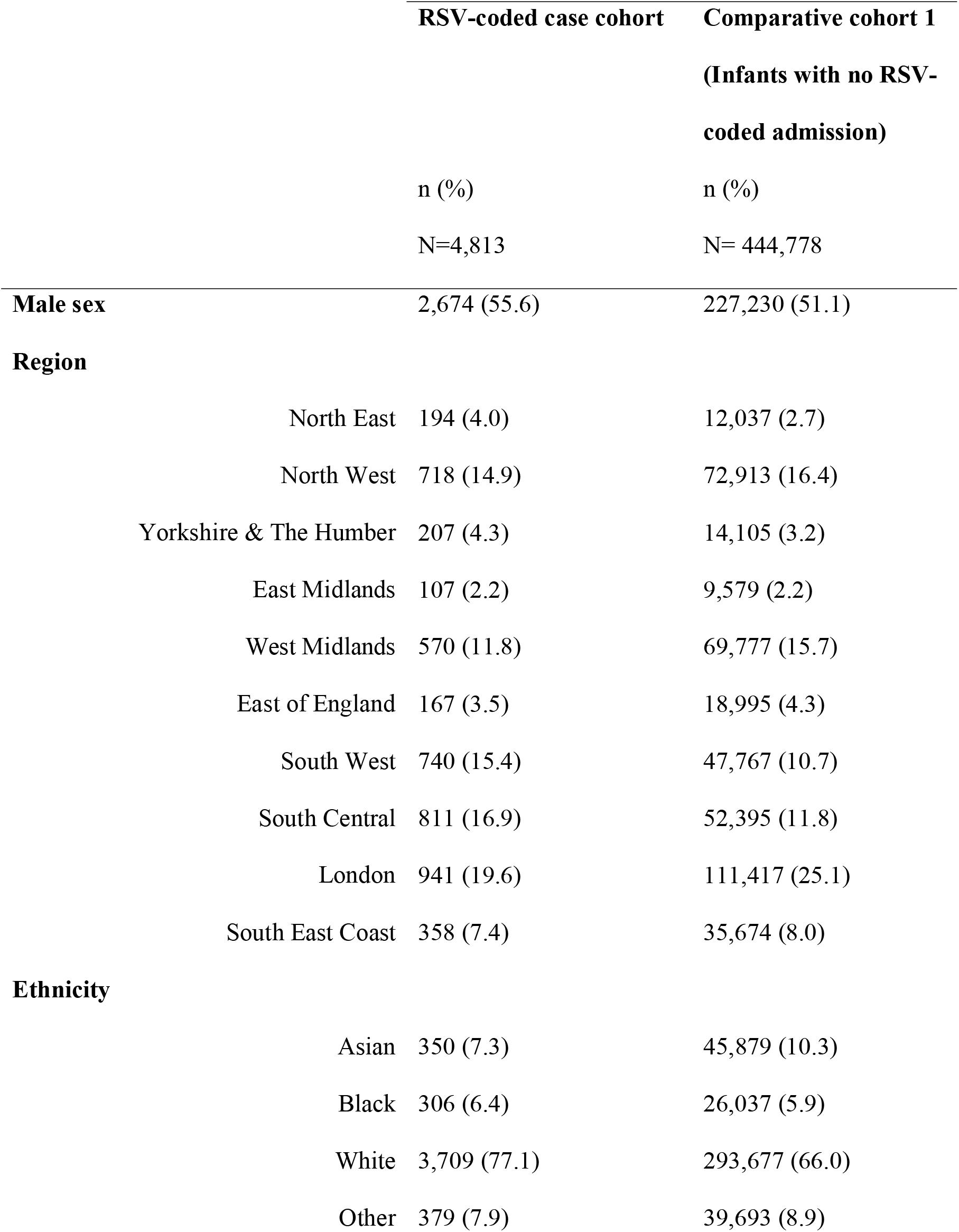
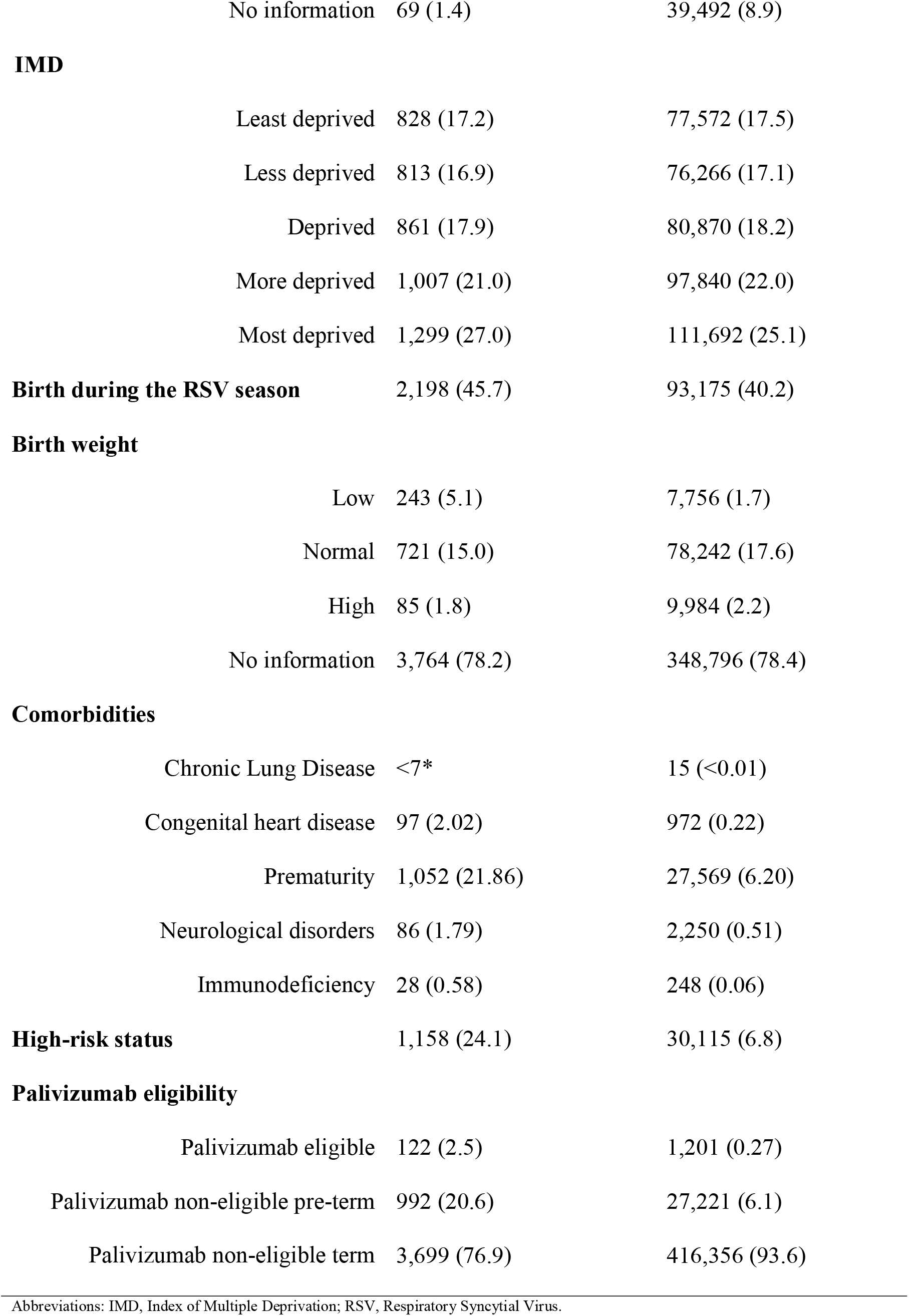
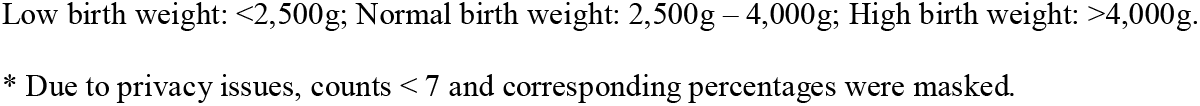
Sociodemographic and Clinical Characteristics of Infants with an RSV-coded Hospital Admission up to 24 Months of Age (RSV-coded Case Cohort) and of Infants with no RSV Admission (Comparative Cohort 1).

A similar analysis was performed for bronchiolitis-coded, RTI-coded and RSV-predicted admissions producing similar results (See Table, Supplemental Digital Content 1).

Additionally, there was little difference in baseline sociodemographic and clinical characteristics between infants born in and before the RSV season for the RSV-coded case cohort (See Table, Supplemental Digital Content 2).

### Rate of Admissions

Among the birth cohort, there was a total of 6,013 RSV-coded admissions (6.7 per 1,000 person-years) up to 24 months of age and, of these, 5,218 (11.6 per 1,000 person-years) occurred up to 12 months of age. The algorithm estimated that 15,457 RSV-predicted admissions (34.4 per 1,000 person-years) occurred up to 12 months of age, representing an additional 10,239 admissions and a 3-fold higher rate. Among the birth cohort, there was a total of 31,754 bronchiolitis-coded admissions (35.3 per 1,000 person-years) and 84,115 RTI- coded admissions (93.6 per 1,000 person-years) up to 24 months of age (Table 2). All types of admissions had a slightly higher rate among infants born during the RSV season compared to infants born before the RSV season (6.0 vs. 7.7 RSV-coded admissions per 1,000 infants born before vs. during the RSV season, respectively) (Table 2).

**Table 2.** Total Number of RSV, Bronchiolitis, any RTI and RSV-Predicted Admissions up to 24 and 12 Months of Life and Respective Rates.

|  | First 24 months of life |  |  |  |  |  | First 12 months of life |  |  |  |  |  |
| --- | --- | --- | --- | --- | --- | --- | --- | --- | --- | --- | --- | --- |
|  | Total |  | Before RSV |  | During RSV |  | Total |  | Before RSV |  | During RSV |  |
|  |  |  | Season Birth |  | Season Birth |  |  |  | Season Birth |  | Season Birth |  |
|  | Number of admissions | Rate | Number of admissions | Rate | Number of admissions | Rate | Number of admissions | Rate | Number of admissions | Rate | Number of admissions | Rate |
| **RSV-coded** | 6,013 | 6.7 | 3,243 | 6.0 | 2,770 | 7.7 | 5,218 | 11.6 | 2,790 | 10.4 | 2,428 | 13.4 |
| **Bronchiolitis-coded** | 31,754 | 35.3 | 18,184 | 33.9 | 13,570 | 37.5 | 28,596 | 63.6 | 16,572 | 61.7 | 12,024 | 66.5 |
| **RTI-coded** | 84,115 | 93.6 | 49,467 | 92.1 | 34,648 | 95.9 | 51,521 | 114.6 | 30,344 | 113.0 | 21,177 | 117.2 |
| **RSV-predicted\*** | - | - | - | - | - | - | 15,457 | 34.4 | 9,181 | 34.2 | 6,276 | 34.7 |

Abbreviations: RSV, Respiratory Syncytial Virus; RTI, Respiratory Tract Infection.
Rate described per 1,000 person-years at risk.
Before Season Birth: birth between 1<sup>st</sup> March to 30<sup>th</sup> September; During the season birth: birth between 1<sup>st</sup> October to 28<sup>th</sup> / 29<sup>th</sup> February.
\*The algorithm was developed for infants up to 12 months of age, and so can only be used in that age group.

### Additional RSV Burden

During the study period, 22.3% and 16.6% of all primary care consultations coded as RTI, and 27.4% and 25.2% of all hospital admissions coded as unspecified LRTI, among infants born before and during the RSV season, respectively, were estimated to be due to RSV (Table 3). The proportions of URTI admissions due to RSV were estimated to be 4.6% and 8.6% among infants born before and during the RSV season, respectively.

**Table 3.** Proportion of Total Primary Care Consultations, and Unspecified LRTI and URTI Hospital Admissions that Were Estimated to Be Due to Each Pathogen, Stratified by Infants Born Before and During the RSV Season.

|  | RTI Primary Care Consultations |  | Unspecified LRTI Hospital Admissions |  | Unspecified URTI Hospital Admissions |  |
| --- | --- | --- | --- | --- | --- | --- |
|  | Before RSV | During RSV | Before RSV | During RSV | Before RSV | During RSV |
|  | Season Birth | Season Birth | Season Birth | Season Birth | Season Birth | Season Birth |
| Pathogen, % (CI) |  |  |  |  |  |  |
| RSV | 22.3 (19.4, 25.2) | 16.6 (14.3, 18.9) | 27.4 (24.3, 30.5) | 25.2 (21.7, 28.6) | 4.6 (1.4, 7.9) | 8.6 (6.1, 11.1) |
| Influenza | 12.7 (9.6, 15.8) | 7.2 (4.7, 9.7) | 7.4 (4.1, 10.7) | 4.6 (0.9, 8.3) | 8.6 (5.7, 11.5) | 10.0 (7.4, 12.5) |
| Parainfluenza | 27.7 (17.4, 37.9) | 34.9 (26.7, 43.0) | 19.3 (8.3, 30.4) | 25.9 (13.5, 38.2) | 9.7 (0.0, 19.5) | 15.6 (7.3, 23.8) |
| Rhinovirus | - | - | - | - | 3.2 (-0.4, 6.9) | - |
| Haemophilus influenzae | - | - | - | - | 2.7 (-0.6, 6.0) | 2.8 (-0.1, 5.6) |

Abbreviations: LRTI, Lower Respiratory Tract Infection; RSV, Respiratory Syncytial Virus; URTI, Upper Respiratory Tract Infection
Pathogens were selected for inclusion using a backward stepwise regression based on the Akaike Information Criterion using the model with the lowest Akaike Information Criterion at each step.
Before Season Birth: birth between 1<sup>st</sup> March to 30<sup>th</sup> September; During the season birth: birth between 1<sup>st</sup> October to 28<sup>th</sup> / 29<sup>th</sup> February.

During the RSV season peak, more than 60% of unspecified LRTI hospital admissions were estimated to be due to RSV (Figure 2). This figure was higher than the peak of the curves of the other pathogens (influenza and parainfluenza). Across the whole RSV season (Oct – Feb), RSV stands out from the other pathogens in the model. For primary care consultations coded as RTI, more than 60% and 50% were due to RSV in the RSV season peak, among infants born before and during season, respectively (Figure 2). The proportion of unspecified URTI hospital admissions estimated to be due to RSV reached 20% and above 30% in the RSV season peak, for infants born before and during season, respectively (Figure, Supplemental Digital Content 3). More pathogens were included in the model (RSV, influenza, parainfluenza, h. influenza and rhinovirus) and the difference in height between the RSV curve and the other pathogens’ curves was not so evident as for unspecified LRTI admissions and RTI primary care consultations.

**Figure 2.**
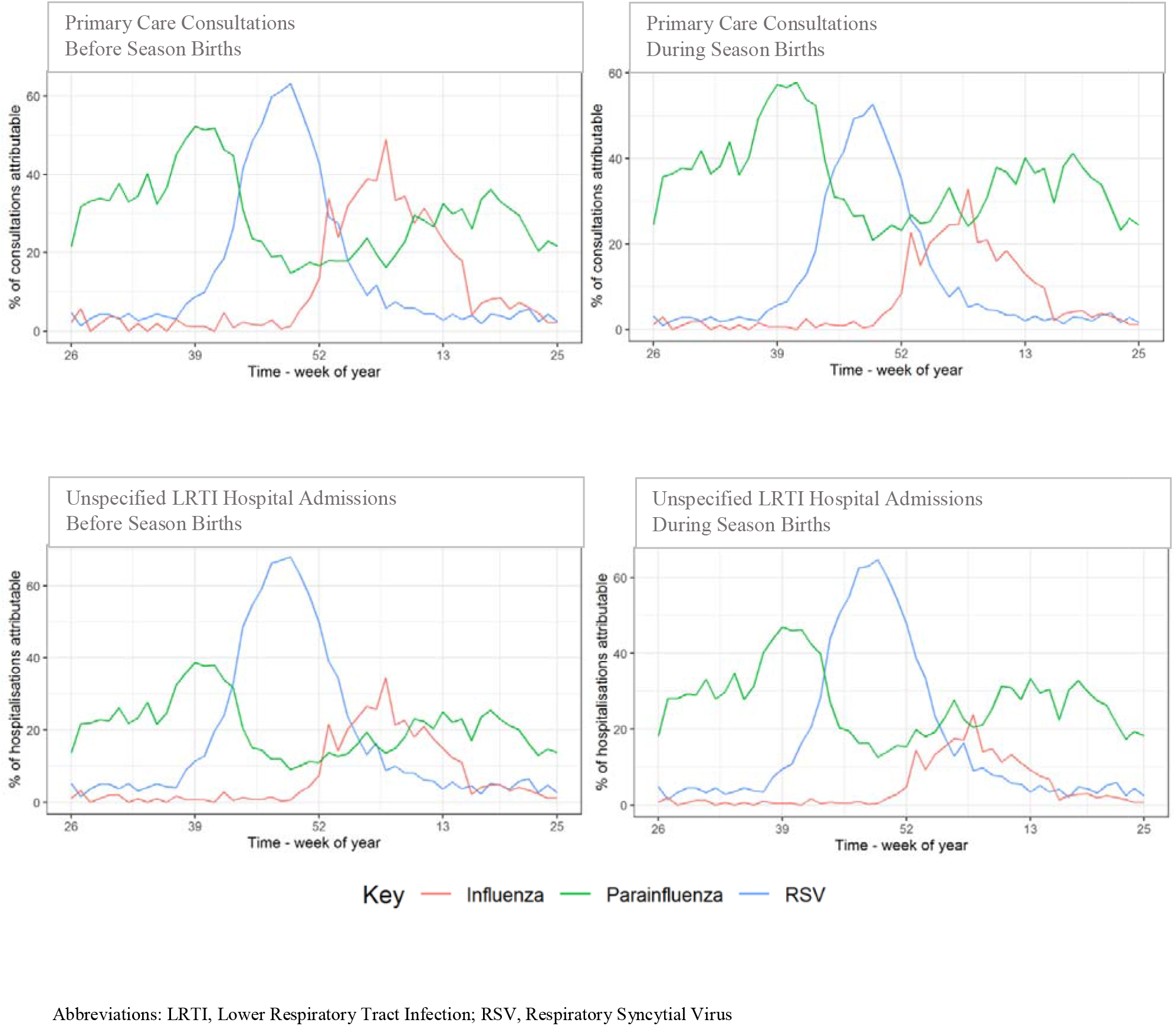
Proportion of Total Primary Care Consultations (1) and Unspecified LRTI (2) Hospital Admissions that Were Estimated to Be Due to Each Pathogen, Stratified by Infants born Before (A) and During (B) the RSV Season, across the Calendar Year.

## Discussion

To our knowledge, this is the first comprehensive analysis of RSV admission rates and patients’ characteristics in England, using the latest pre-pandemic data, a birth cohort design, and different approaches to identify RSV. Using RSV specific codes, we found an incidence of 1.1% during the first 24 months of life, and a rate of 6.7 per 1,000 person-years, but a higher rate during the first 12 months of life - 11.6 per 1,000 person-years. When using an algorithm to predict RSV admissions, both the incidence and rate were ∼3 times higher than coded admissions only. Most infants hospitalized with RSV before the age of 24 months were term healthy infants (>74%) born before the RSV season (>54%). Infants born during the RSV season have similar baseline characteristics and a marginally higher rate of RSV admissions compared to those born before season.

Our findings in the English healthcare setting are consistent with previous literature, which shows that RSV causes considerable burden of disease in infants worldwide.^1^ While the majority of RSV related deaths occurs in low- and middle-income countries, there is still a substantial RSV related healthcare burden in high income countries.^6,19^ In England, only 7% of all RTI admissions have a pathogen-specific diagnosis, meaning that the vast majority of RTI admissions have no causal pathogen coded^6^, likely leading to an underestimation of the true burden of RSV in secondary care, when using RSV-coded admissions only. To overcome this, we applied an algorithm, based on Reeves et al.^9^, to identify RTI admissions that were due to RSV, estimating a rate ∼3 times higher than when using coded admissions only. Additionally, to understand the total burden of RSV, through statistical modeling, we estimated that, in infants up to 24 months of age, 22% and 17% of RTI primary care consultations and 27% and 25% of unspecified LRTI hospital visits were due to RSV, for infants born before and during the RSV season, respectively. During the RSV season, these proportions can go up to around 60%. These results are in line with previous literature^8^ and point to an underestimation of the true burden of RSV infection in England, highlighting the impact of RSV on population health.

Infants in the RSV-coded cohort were more likely to be male, more deprived, born in the south-west/south-central regions of England and White than those in the comparative cohort. The deprivation findings, are in line with previous studies showing a higher incidence of respiratory infections in deprived areas^20,21^, and underline an opportunity to reduce health inequalities through a national level immunization strategy covering all infants. Low birth weight and comorbidities, including prematurity, were more frequent among those with an RSV admission, which is supported by previous literature.^22^ However, the majority of infants hospitalized with RSV were born at term and were previously healthy, similar to findings from other studies.^23–25^ While it is important to identify infants at greatest risk of severe RSV infection in order to determine those who could benefit most from interventions aiming to either prevent infection or reduce disease severity, prevention strategies targeting only high-risk infants will have a limited effect at the population level, in particular on the total burden of RSV infection.^23^ Results from the bronchiolitis-coded, RTI-coded and RSV-predicted cohorts corroborate the results from the RSV-coded cohort.

Another well recognized risk factor for RSV hospitalization is birth during the RSV season.^15,26^ This was confirmed in our research where infants who had an RSV admission were more likely to be born during the RSV season (October to February) than their peers with no admission. However, over half of the infants hospitalized with RSV or bronchiolitis or predicted to be hospitalized with RSV were born before the RSV season, reflecting the greater size of the out of season cohort. As previously observed,^27^ when stratifying by before and during the season birth, baseline sociodemographic and clinical characteristics were similar across both groups and rates of RSV admission were marginally higher in the group born during the season. While infants born during the RSV season were more likely to be hospitalized with an RSV infection due to exposure to RSV at a younger age, combined with lower levels of RSV antibodies in the maternal population early in the RSV season^15^, the limited differences between those born before and during the RSV season found in the current study stress that preventative efforts should include all infants entering their first season of RSV.

The main strength of this study is the birth cohort design with the power to accurately estimate the admission rates of RSV in England. The birth cohort design applied to a large sample size representative of the English population allowed us to define the exact “time at risk” to which each infant contributed to the analysis, adding to the literature precise and up-to-date estimates of the incidence and rates of hospitalizations due to RSV in England. Yet, we know RSV is poorly coded and to overcome this limitation we built three additional case cohorts, whose results corroborate findings from the RSV-coded case cohort, and we also used a statistical modelling approach to ascertain the proportion of unspecified admissions that were estimated to be due to RSV. A limitation of the RSV-predicted case cohort is that the algorithm was designed for infants up to 12 months of age, whereas the other case cohorts account for cases up to 24 months of age. Another limitation of the algorithm is that a fixed-coefficient logistic model was applied to different data, and consequently the prediction of RSV admissions may not be as accurate as in the original data.^9^ We are aware that palivizumab prophylaxis was recommended for some high-risk infants, however, the identification of those who actually received palivizumab was not possible. Lastly, at the time the protocol was developed, there was uncertainty around the effects of the COVID-19 pandemic on RSV activity and for that reason we decided to include the latest data up to the start of the pandemic.

## Conclusion

Our study has found that, in England, one in 91 infants have an RSV-coded admission, which is likely underestimated by around 3-fold. In absolute terms, most of these are healthy infants born at term before the RSV season, confirming the high burden of RSV and supporting an immunization strategy targeted at protecting all infants entering their first RSV season. This is the optimum strategy to protect at the population level and decrease the total burden of RSV infection on infants, their families, and the NHS and contribute to reducing health inequalities with regards to RSV hospitalization.

## Data Availability

All data produced in the present study are available upon reasonable request to the authors

## Acknowledgments

We thank Lori Cirneanu and Jessica Lundbom for their support with the statistical analysis. We thank Rachel Reeves for her expertise and assistance in the protocol development and Nuria Martinez-Alier for her clinical input. We also thank Craig Davidson and Mersha Chetty for their support in results interpretation. This study is based in part on data from the Clinical Practice Research Datalink obtained under licence from the UK Medicines and Healthcare products Regulatory Agency. The data is provided by patients and collected by the NHS as part of their care and support. The interpretation and conclusions contained in this study are those of the authors alone. Copyright © (2021), re-used with the permission of The Health & Social Care Information Centre. All rights reserved.

## Ethics approval

This retrospective study using secondary data was conducted in accordance with the ethical standards of the institutional and national research committees and with the Declaration of Helsinki. The Medicines and Healthcare products Regulatory Agency (MHRA) Independent Scientific Advisory Committee (ISAC) approved this study.

## Consent to participate

Informed consent was unnecessary as the study used only electronic health medical records, which had been de-identified prior to data access.

## Supplemental Digital Content

**Supplemental Digital Content 1.**
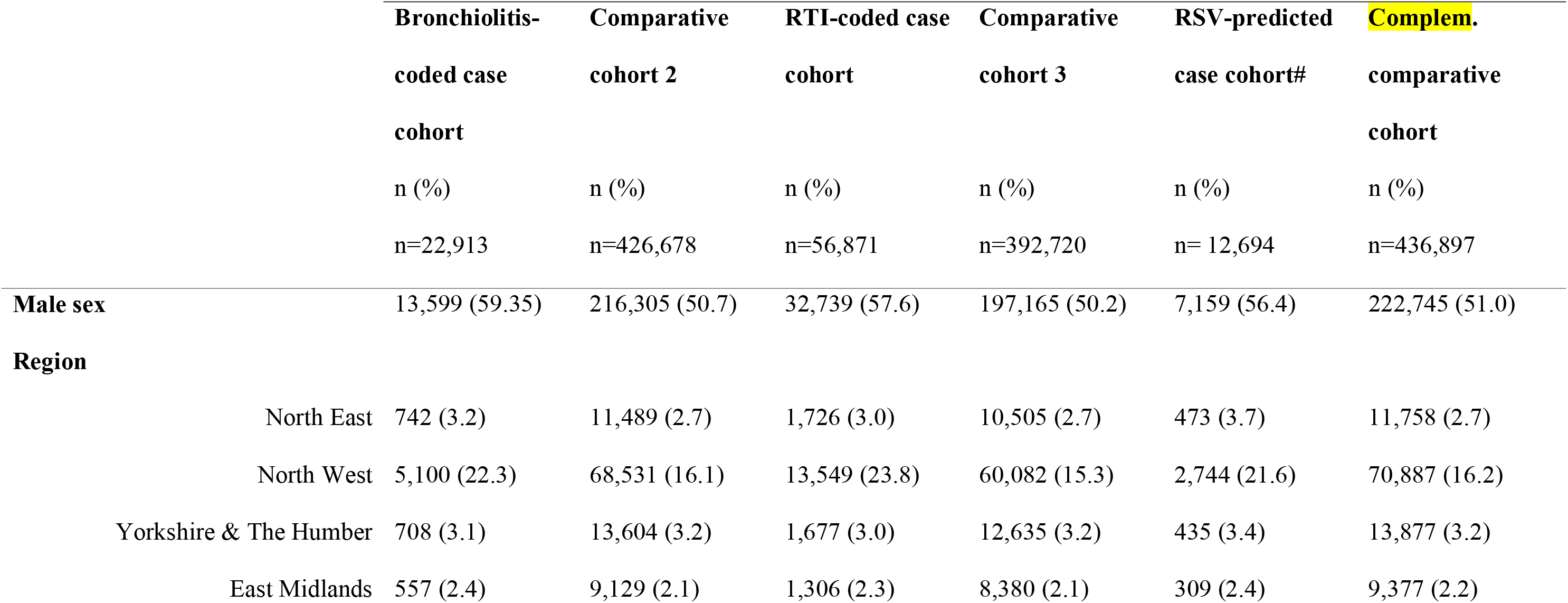
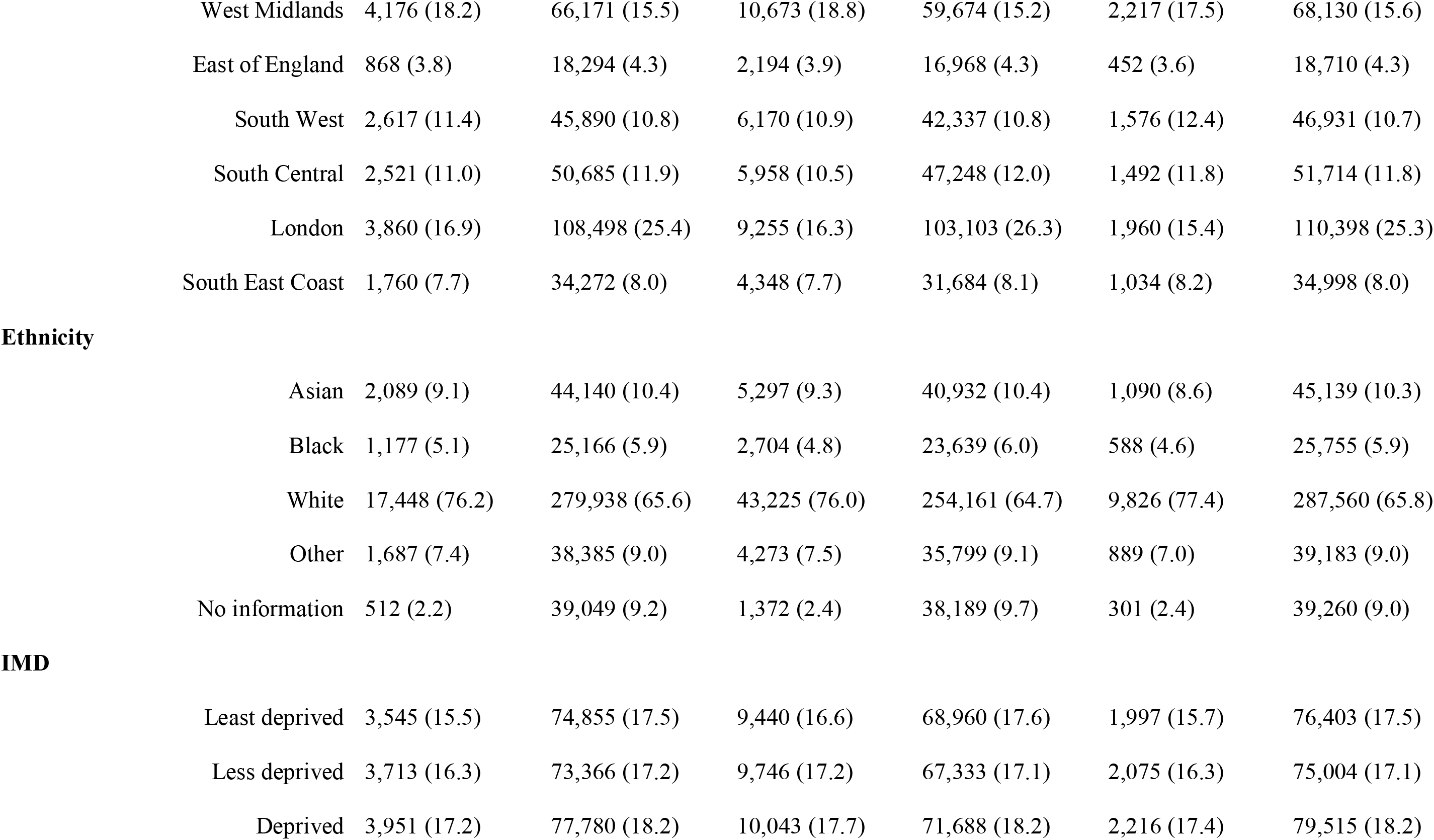
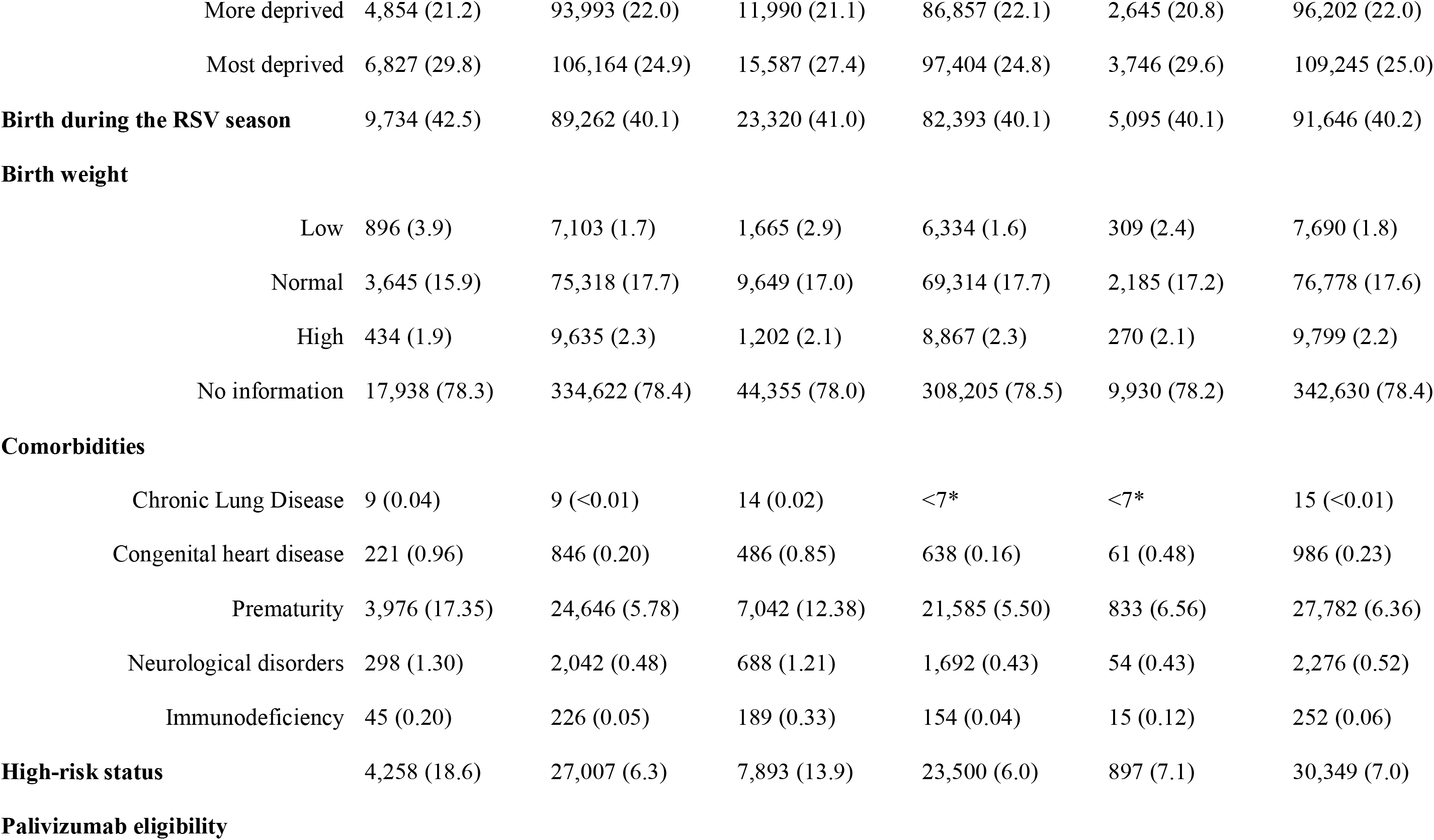
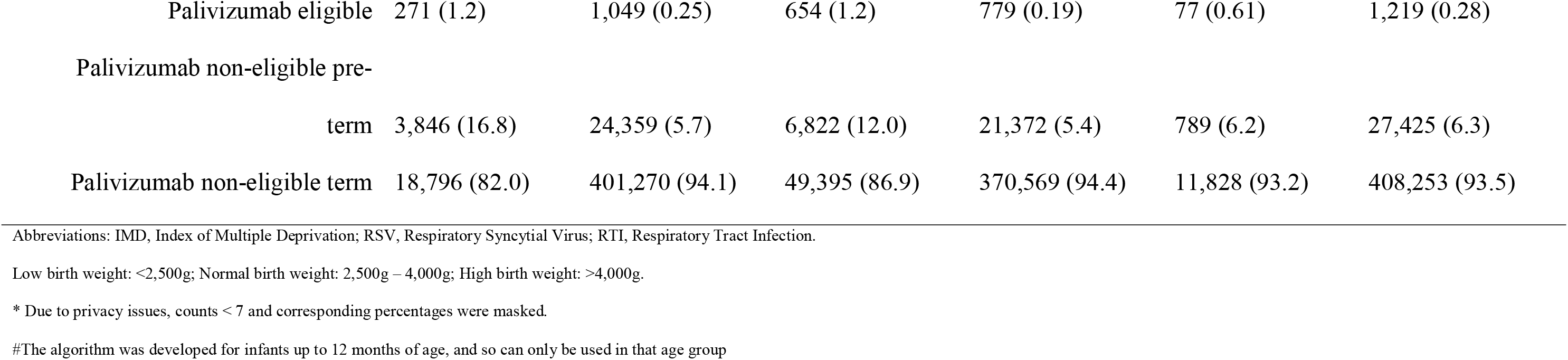
Sociodemographic and Clinical Characteristics of Infants with a bronchiolitis-coded, and RTI-coded Hospital Admission up to 24 Months of Age and RSV-predicted Hospital Admission up to 12 Months of Age (Case Cohorts) and of Infants with no Admission (Comparative Cohorts).

**Supplemental Digital Content 2.**
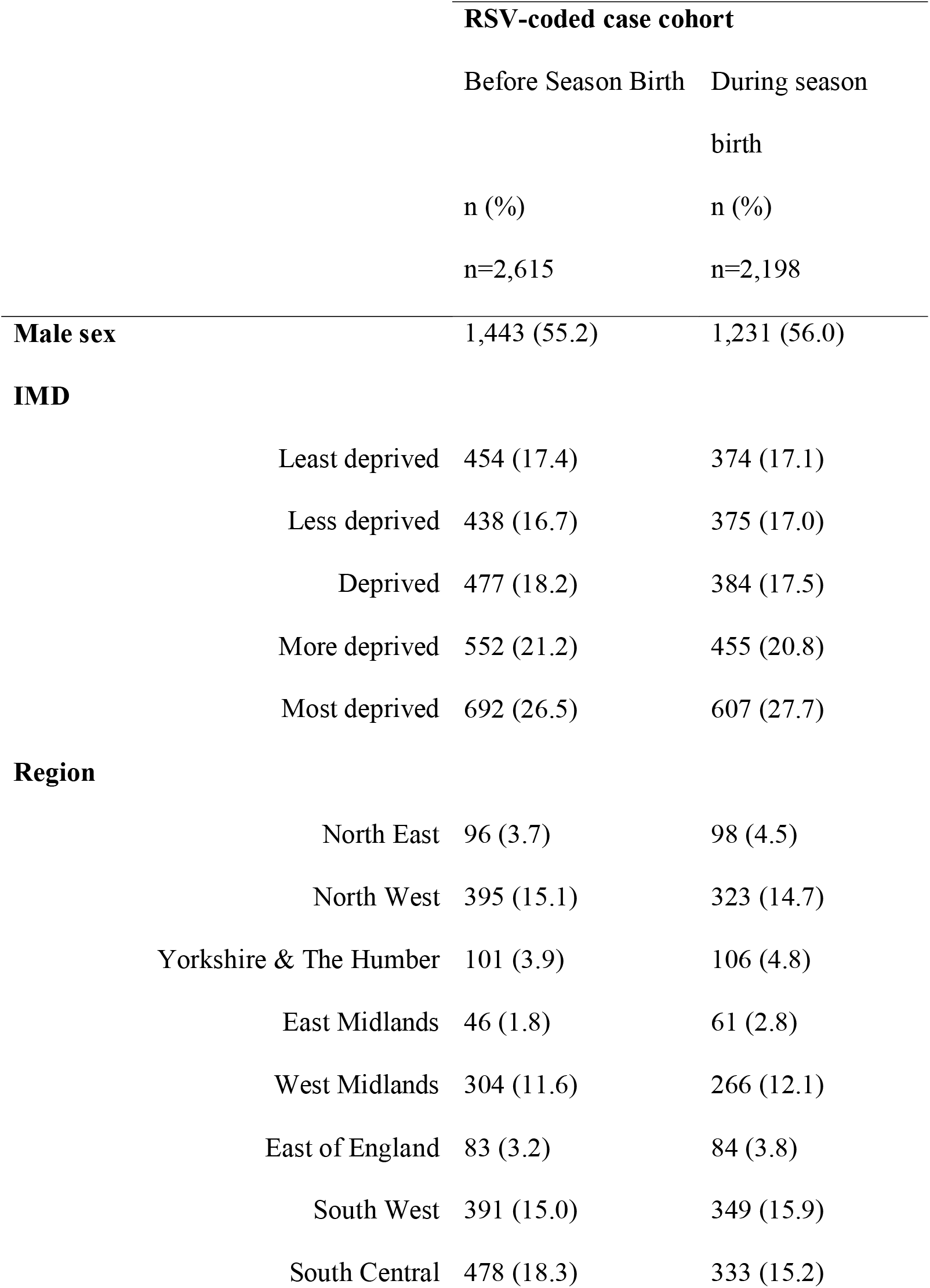
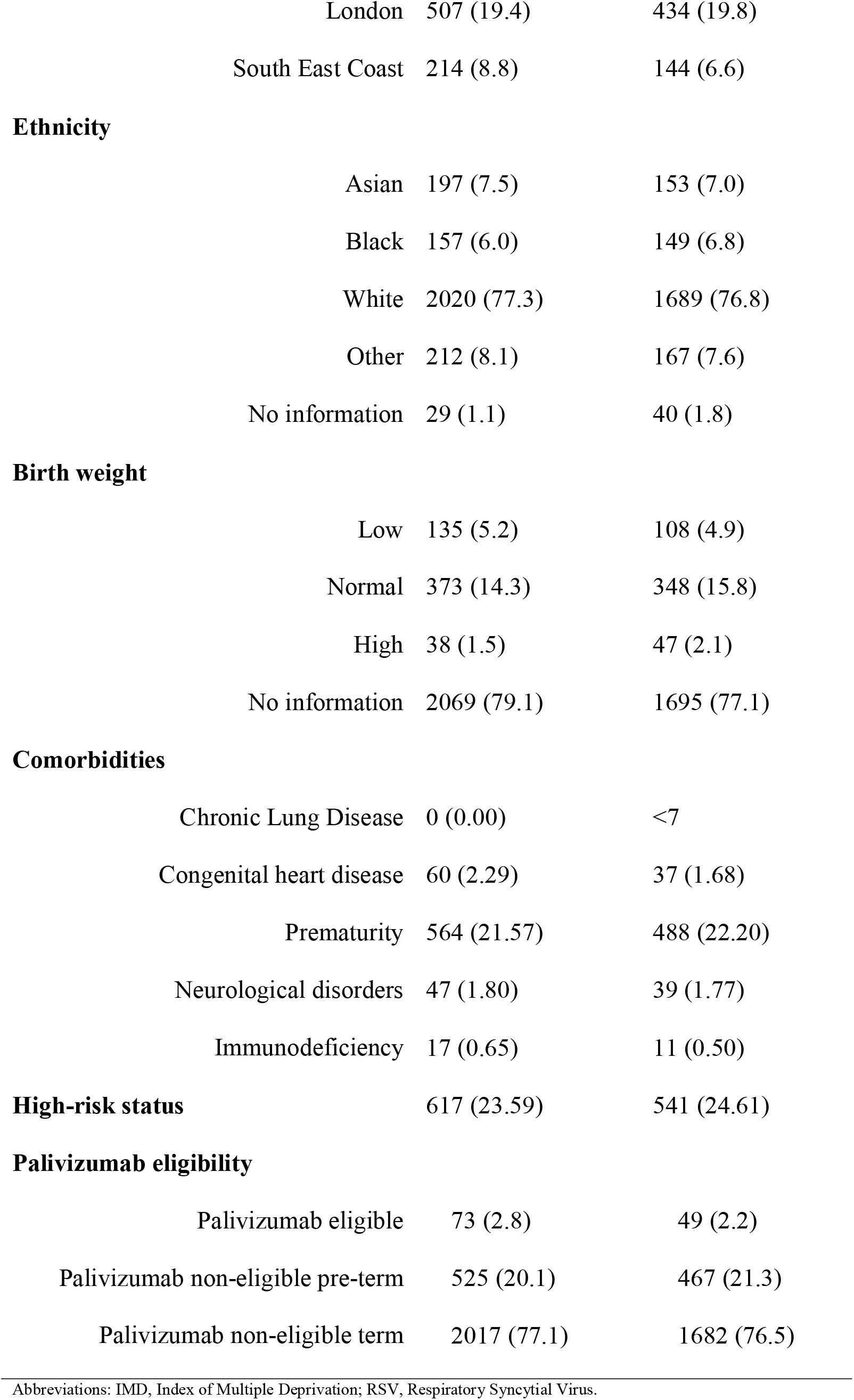
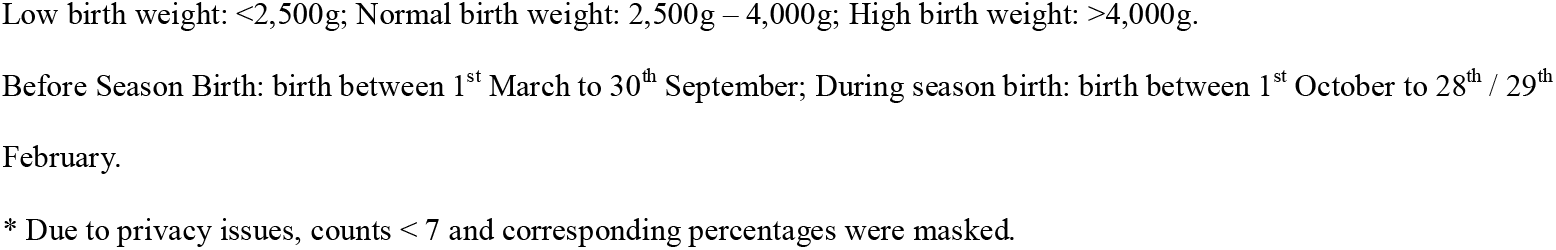
Sociodemographic and Clinical Characteristics of Infants with an RSV-coded Hospital Admission up to 24 Months of Age (RSV-coded Case Cohort), Stratified by Infants Born Before and During the RSV Season.

**Supplemental Digital Content 3.**
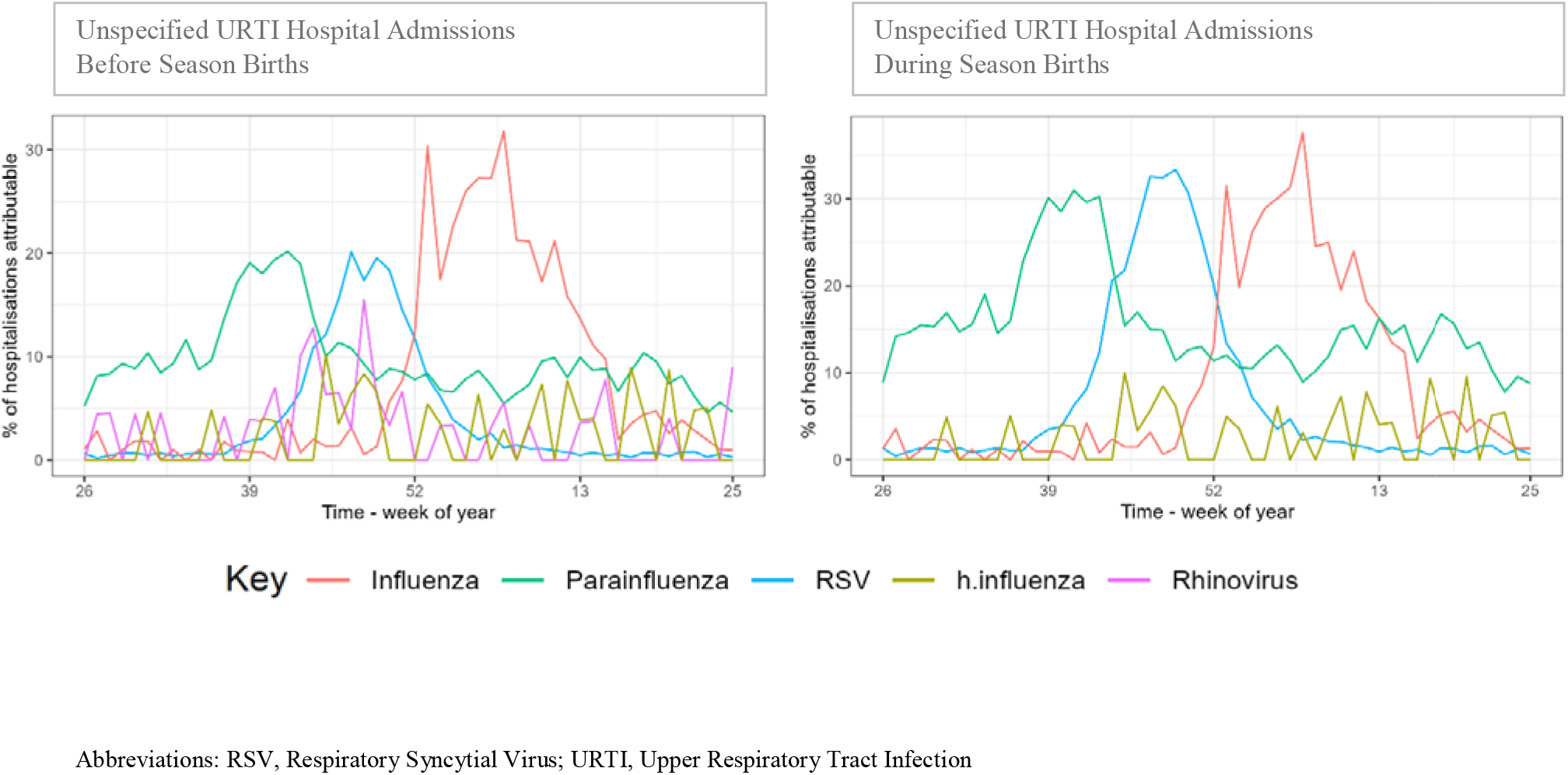
Proportion of Total Unspecified URTI Hospital Admissions that Were Estimated to Be Due to Each Pathogen, Stratified by Infants Born Before and During the RSV Season, across the Calendar Year.

## References

Li Y, Wang X, Blau DM, et al. Global, regional, and national disease burden estimates of acute lower respiratory infections due to respiratory syncytial virus in children younger than 5 years in 2019: a systematic analysis. Lancet. 2022;399(10340):2047– 2064.

Collins PL, Fearns R, Graham BS. Respiratory syncytial virus: virology, reverse genetics, and pathogenesis of disease. Curr Top Microbiol Immunol. 2013;372:3–38.

Young M, Smitherman L. Socioeconomic Impact of RSV Hospitalization. Infect Dis Ther. 2021;10(Suppl 1):35–45.

Blanken MO, Rovers MM, Molenaar JM, et al. Respiratory Syncytial Virus and Recurrent Wheeze in Healthy Preterm Infants. New England Journal of Medicine. 2013;368(19):1791–1799.

Fauroux B, Simões EAF, Checchia PA, et al. The Burden and Long-term Respiratory Morbidity Associated with Respiratory Syncytial Virus Infection in Early Childhood. Infect Dis Ther. 2017;6(2):173–197.

Reeves RM, van Wijhe M, Tong S, et al. Respiratory Syncytial Virus-Associated Hospital Admissions in Children Younger Than 5 Years in 7 European Countries Using Routinely Collected Datasets. The Journal of Infectious Diseases. 2020;222(Supplement_7):S599–S605.

Pitman RJ, Melegaro A, Gelb D, et al. Assessing the burden of influenza and other respiratory infections in England and Wales. J Infect. 2007;54(6):530–538.

Reeves RM, Hardelid P, Gilbert R, et al. Estimating the burden of respiratory syncytial virus (RSV) on respiratory hospital admissions in children less than five years of age in England, 2007-2012. Influenza Other Respir Viruses. 2017;11(2):122–129.

Reeves RM, Hardelid P, Panagiotopoulos N, et al. Burden of hospital admissions caused by respiratory syncytial virus (RSV) in infants in England: A data linkage modelling study. J Infect. 2019;78(6):468–475.

Chung A, Reeves RM, Nair H, et al. Hospital Admission Trends for Bronchiolitis in Scotland, 2001–2016: A National Retrospective Observational Study. The Journal of Infectious Diseases. 2020;222(Supplement_7):S592–S598.

Schindeler SK, Muscatello DJ, Ferson MJ, et al. Evaluation of alternative respiratory syndromes for specific syndromic surveillance of influenza and respiratory syncytial virus: a time series analysis. BMC Infect Dis. 2009;9:190.

Cai W, Tolksdorf K, Hirve S, et al. Evaluation of using ICD-10 code data for respiratory syncytial virus surveillance. Influenza Other Respir Viruses. 2020;14(6):630–637.

Pignotti MS, Carmela Leo M, Pugi A, et al. Consensus conference on the appropriateness of palivizumab prophylaxis in respiratory syncytial virus disease. Pediatr Pulmonol. 2016;51(10):1088–1096.

American Academy of Pediatrics Committee on Infectious Diseases, American Academy of Pediatrics Bronchiolitis Guidelines Committee. Updated guidance for palivizumab prophylaxis among infants and young children at increased risk of hospitalization for respiratory syncytial virus infection. Pediatrics. 2014;134(2):415–420.

Reeves RM, Hardelid P, Gilbert R, et al. Epidemiology of laboratory-confirmed respiratory syncytial virus infection in young children in England, 2010-2014: the importance of birth month. Epidemiol Infect. 2016;144(10):2049–2056.

Clinical Practice Research Datalink. (2022). CPRD Aurum November 2022 (Version 2022.11.001) [Data Set]. Clinical Practice Research Datalink. Https://Doi.Org/10.48329/Cbh0-4e76.

Respiratory syncytial virus: the green book, chapter 27a. https://assets.publishing.service.gov.uk/government/uploads/system/uploads/attachment_data/file/458469/Green_Book_Chapter_27a_v2_0W.PDF. Accessed August 8, 2022.

Müller-Pebody B, Edmunds WJ, Zambon MC, et al. Contribution of RSV to bronchiolitis and pneumonia-associated hospitalizations in English children, April 1995- March 1998. Epidemiol Infect. 2002;129(1):99–106.

Wildenbeest JG, Billard M-N, Zuurbier RP, et al. The burden of respiratory syncytial virus in healthy term-born infants in Europe: a prospective birth cohort study. The Lancet Respiratory Medicine. 2022;0(0).

Zheng Z, Warren JL, Shapiro ED, et al. Estimated incidence of respiratory hospitalizations attributable to RSV infections across age and socioeconomic groups. Pneumonia. 2022;14(1):6.

Chung A, Reeves RM, Nair H, et al. Hospital Admission Trends for Bronchiolitis in Scotland, 2001-2016: A National Retrospective Observational Study. J Infect Dis. 2020;222(Suppl 7):S592–S598.

Shi T, Balsells E, Wastnedge E, et al. Risk factors for respiratory syncytial virus associated with acute lower respiratory infection in children under five years: Systematic review and meta-analysis. J Glob Health. 2015;5(2):020416.

Hall CB, Weinberg GA, Iwane MK, et al. The burden of respiratory syncytial virus infection in young children. N Engl J Med. 2009;360(6):588–598.

Bandeira T, Carmo M, Lopes H, et al. Burden and severity of children’s hospitalizations by respiratory syncytial virus in Portugal, 2015–2018. Influenza and Other Respiratory Viruses. 2023;17(1):e13066.

Martinón-Torres F, Carmo M, Platero L, et al. Clinical and economic burden of respiratory syncytial virus in Spanish children: the BARI study. BMC Infect Dis. 2022;22(1):759.

Sommer C, Resch B, Simões EAF. Risk Factors for Severe Respiratory Syncytial Virus Lower Respiratory Tract Infection. Open Microbiol J. 2011;5:144–154.

Demont C, Petrica N, Bardoulat I, et al. Economic and disease burden of RSV-associated hospitalizations in young children in France, from 2010 through 2018. BMC Infect Dis. 2021;21(1):730.

